# AI-based deformable hippocampal mesh reflects hippocampal morphological characteristics in relation to cognition in healthy older adults

**DOI:** 10.1101/2024.10.28.24316272

**Authors:** Wonjung Park, Maria del C. Valdés Hernández, Jaeil Kim, Susana Muñoz Maniega, Fraser Sneden, Karen J. Ferguson, Mark E. Bastin, Joanna M. Wardlaw, Simon R. Cox, Jinah Park

## Abstract

Magnetic resonance imaging (MRI)-derived hippocampus measurements have been associated with different cognitive domains. The knowledge of hippocampal structural deformations as we age has contributed to our understanding of the overall aging process. Different morphological hippocampal shape analysis methods have been developed, but it is unclear how their principles relate and how consistent are the published results in relation to cognition in the normal elderly in the light of the new deep-learning-based (DL) state-of-the-art modeling methods. We compared results from analysing the hippocampal morphology using manually-generated binary masks and a Laplacian-based deformation shape analysis method, with those resulting from analysing SynthSeg-generated hippocampal binary masks using a DL method based on the PointNet architecture, in relation to different cognitive domains. Whilst most previously reported statistically significant associations were also replicated, differences were also observed due to 1) differences in the binary masks and 2) differences in sensitivity between the methods. Differences in the template mesh, number of vertices of the template mesh, and their distribution did not impact the results.

**Highlights:**

- Newdeep-learning-based hippocampal 3D-shape modeling method replicates hippocampal shape reported associations with cognition
- New deep-learning-based hippocampal 3D-shape modeling method has increased sensitivity than a conventional Lapalcian-based deformation method
- Accuracy in hippocampal binary masks is crucial in the AI-based shape modeling method

## 1. Introduction

The hippocampus is probably the most studied brain structure given its role in cognitive processes. These range from memory (Bird and Burgess, 2008) to processing speed (Papp et al., 2014) and fluid intelligence (Reuben et al., 2011). In-vivo measurements of hippocampi total and subfields’ volumes using magnetic resonance imaging (MRI) (Voineskos et al., 2015), sometimes combined with measurements of quantitative parameters (e.g., T1, fractional anisotropy, mean diffusivity) (Anblagan et al., 2018), symmetry between hippocampi in the left vs. right hemispheres (Woolard and Heckers, 2012), and analyses of their shape (Hernández et al., 2017), (Scher et al., 2007), have advanced our understanding on hippocampi role with cognition (Maruszak and Thuret, 2014) and, most importantly, the relationship between how the brain ages and how this aging process looks like in terms of functional performance and overall health indicators.

Importantly, hippocampal shape has been considered of utmost relevance to the study of Alzheimer’s disease (AD) and the path leading to the onset of this disease. An atrophy scale of the middle temporal lobe with high diagnostic value particularly considers the height of the hippocampi (Scheltens et al., 1992). Differential hippocampi subfield atrophy, how shape may be affected, and whether this may be a preclinical indicator for developing AD have been also areas of active research (Mueller et al., 2010), (Csernansky et al., 2005). But the literature on the theme is not free from contradictory results or results that haven’t been replicated, even in samples with similar clinical and demographic characteristics. Difficulties and discrepancies in establishing the limits of the hippocampi subfields given by inter-subject variability (Amunts et al., 2005) and differences in imaging protocols (Van Strien et al., 2012) have been previously discussed. Differences in the precision of computational methods segmenting the hippocampi to give absolute volumes have also been documented (Hasboun et al., 1996), (Barnes et al., 2008), (Mulder et al., 2014). However, differences in hippocampal shape deformations in relation to cognition between studies have been largely attributed to differences on brain aging trajectories (Hernández et al., 2017) rather than to differences in the methods and references used to derive these shape deformations.

### 1.1. Related works using traditional shape modeling

Several methods that manipulate and combine geometrical shapes to describe a 3D object have been developed and used to analyze hippocampal shape. Some of these are: adaptive-focus deformable model of a single hippocampus using attribute vectors and a shape energy function (Shen et al., 2002), medial shape representation (Gerig et al., 2003), spherical harmonic representation (SPHARM) (Shen et al., 2003) (Brechbühler et al., 1995) and their combination (i.e., SPHARM-based medial representation) (Styner et al., 2003), the conversion of binary hippocampal segmentations into their corresponding spherical harmonic description (i.e.,SPHARM) later sampled into a triangulated surface (SPHARM-PDM) (Styner et al., 2006), combination of medial shape surfaces and local width analyzes (Bouix et al., 2005), diffeomorphic maps (Csernansky et al., 2005) including the template-based large deformation diffeomorphic metric mapping (LDDMM) approach (Raamana et al., 2014) subsequently embedded into comprehensive frameworks like the automatic segmentation of hippocampal subfields (ASHS) (Yushkevich et al., 2015), Laplacian-based template surface progressive deformation (Kim et al., 2014) combined with differential geometry operators (Diers et al., 2023), combination of the minimum description length criterion (MDL) with shape re-parameterization using Cauchy kernels (Davies et al., 2003), entropy-based particle systems (Cates et al., 2007), and kernel smoothing of skeletal representations (García-Portugués and Meilán-Vila, 2023).

As these models and modeling methods manipulate geometrical shapes mathematically defined (e.g., triangles, polygons, lines concatenated in a skeleton among others) to represent the hippocampus, and describe its deformities (i.e., with respect to a mean shape) also using geometrical, differential, or geodesic principles, we refer to them throughout as *traditional models* or *non-AI-based models*. These methods achieve the fitting of the hippocampal mesh (i.e., formed by the combination of the aforementioned geometrical shapes) to the binary masks by acting directly on the edges, vertices, and surfaces, or by manipulating the parameters of a deformation modeling method that governs the displacements of these edges, vertices, and surfaces in the meshes.

### 1.2. Related works using AI-based shape modeling

With the advancement of deep learning (DL), shape reconstruction technology that converts regions of interest (RoI) in images or point clouds representations into meshes has gained significant attention. Pixel2Mesh (Wang et al., 2018) generates 3D meshes from single RGB images, predicting the overall 3D surface from highly constrained 2D data. In addition, Point2Mesh (Hanocka et al., 2020) creates 3D meshes from point clouds by learning the self-prior of the target object.

In biomedical research, several methods have been proposed to reconstruct the shape of body organs using deep learning. Voxel2Mesh (Wickramasinghe et al., 2020) introduces a CNN-based approach that directly converts 3D images into liver surfaces. Cortical surfaces have been reconstructed from MRI scans using a graph neural network (GNN)-based architecture (Bongratz et al., 2024), and vertebrae shape has been reconstructed with a differentiable signed distance operator (Kim and Park, 2024). In addition, GNN-based approaches have been applied to reconstruct cardiac shapes from sparse and incomplete point clouds (Chen et al., 2021), and whole-heart shapes from image data (Kong et al., 2021). To our knowledge, there has not yet been a deep learning-based approach specifically for hippocampal shape reconstruction, and neither any adaptation of the existent deep learning-based shape models to analyze its relation to cognition.

In this paper, we address the reconstruction of hippocampal shape using multi-layer perceptron (MLP)-based Point-Net (Qi et al., 2017) architecture with the optimization process from Point2Mesh (Wang et al., 2018). We first generate point clouds from individual hippocampal masks and then convert these point clouds into 3D meshes. As demonstrated by previous shape reconstruction methods (Bongratz et al., 2024; Kong et al., 2021), to ensure point correspondence across all meshes for cross-sectional analysis, we deform a template hippocampal mesh to create individual meshes, ensuring that all meshes share the same vertex connectivity.

### 1.3. Contributions

Results from using traditional methods (i.e., defined as per in Section 1.1) to analyze hippocampal shape deformations in relation to cognition in the elderly, individuals with mild cognitive impairment (MCI), and Alzheimer’s disease (AD) patients, mostly agree in the areas where inwards and outwards deformations with respect to a population template relate to different cognitive functions. For example, Kim and colleagues (Kim et al., 2014) compared the performance of LDDMM, SPHARM-PDM and a Laplacian-based template surface progressive deformation modeling method in discerning the hippocampal regions of significant morphological shape differences between 50 AD patients, 50 individuals with MCI, and 50 cognitively normal individuals from the Alzheimer’s Disease Neuroimaging Initiative (ADNI) database, obtaining similar results from the three methods (Kim et al., 2014). Linear regression models also showed similar patterns in the shape deformations in relation to Mini-Mental State Examination (MMSE) scores.

Nowadays, many AI-based methods are replacing traditional approaches not only in the tasks of image classification and segmentation, but also in the area of 3D shape modeling and analysis. While these AI-based methods offer certain advantages, their outcome for analyzing the hippocampi shape in relation to cognition hasn’t been compared with previously published results from the more traditional shape analysis methods. The advancements of science and technology impose the necessity to re-evaluate our former knowledge to strengthen the grounds in which the new knowledge is based and to better understand their respective strengths and limitations.This is precisely the main aim and contribution of this study.

Specific contributions are three fold: 1) We present an AI-based hippocampal deformation model that optimizes the fitting of the vertices of the template mesh to a point cloud constructed from the hippocampal binary masks using a deep neural network architecture; 2) we perform a systematic comparison of the Laplacian-based framework proposed by Kim and colleagues (Kim et al., 2014) (Kim et al., 2019) with our new AI-based framework; and 3) we

evaluate the influence of a) template shape, b) mesh vertices distribution (i.e., evenly/uniform vs. non-uniform), c) number of vertices, d) hippocampal segmentation methods (i.e., manually/semi-automatically vs. automatically), and e) shape modeling method, in the analyzes of the hippocampal deformations in a large community-dwelling sample in relation to cognition. We also analyze the causes of the discrepancies where they exist, to draw recommendations on when and how to use each method, and discuss the implications for past and future research.

## 2. Materials and methods

### 2.1. Dataset

Data was provided by The Lothian Birth Cohort 1936 (LBC1936; (Deary et al., 2007)) Study database (https://lothian-birth-cohorts.ed.ac.uk/data-access-collaboration). The LBC1936 study is a large longitudinal study on cognitive aging which recruited community-dwelling cognitively normal individuals born in 1936 in Edinburgh and the Lothians regions in Scotland, who volunteered and gave informed consent to participate in the observational study. Most of them underwent brain magnetic resonance imaging (MRI) in 2008-2010 at mean age 72.6 years old and since every 3 years up to date, and completed the same battery of cognitive tests at each assessment wave. For the present analysis we used imaging and cognitive data from the first MRI scanning wave and the corresponding cognitive testing (i.e., second) wave from the 654 study participants from which we could retrieve both: MRI and complete cognitive data at this wave. The imaging acquisition and processing protocol of the LBC1936 is described in (Wardlaw et al., 2011).

#### 2.1.1. MRI derived data: hippocampal binary masks

This study uses hippocampal binary masks generated by two different methods. The first is semi-automatic, and consists in manually editing the output from an in-house pipeline that uses as input the T1-weighted MRI, and the following combination of tools from the FMRIB Software Library, version 4.1 (FSL; https://fsl.fmrib.ox.ac.uk/fsl/docs/#/): SUSAN (https://users.fmrib.ox.ac.uk/~steve/susan/susan/node18.html) with a filter kernel of [-1 3 3 1], the first-flirt script adapted to use an age-relevant template ((Armitage et al., 2024); https://doi.org/10.7488/ds/7763), and FIRST((Patenaude et al., 2011); https://fsl.fmrib.ox.ac.uk/fsl/docs/#/structural/first) for generating the binary masks of left and right hippocampi. If the hippocampal masks from the automatic pipeline were incorrect, these were generated manually using Analyze 10.0 (https://analyzedirect.com/) following published guidelines (MacLullich et al., 2002), (Kim et al., 2019). The second method is fully automatic, and uses SynthSeg (Billot et al., 2023), a DL method. For our experiments we excluded 53 cases due to poor image quality leading to unusable hippocampal masks, leaving a total of 601 study participants’ data used in our experiments.

#### 2.1.2. Cognitive data

This study uses results from selected subtests from the Wechsler Adult Intelligence Scale (WAIS-III; (Wechsler, 1997)), the Wechsler Memory Scale III (WMS III; (Wechsler, 1998)), measures of simple and 4-choice reaction time and inspection time (Deary et al., 2007), all grouped into three cognitive domains: memory, information processing speed, and general fluid intelligence, by using principal component analysis. Thus, we refer throughout to these three latent variables as *g-memory, g-speed* and *g* respectively. These were used previously in (Hernández et al., 2017). The proportion of variances explained by the first component in each of these cognitive domains can be found in the aforementioned publication.

### 2.2. Hippocampal shape analysis using a Laplacian-based template deformation

We use the shape modeling method described in (Kim et al., 2014). The procedure is explained in details in (Kim et al., 2019), and the code is downloadable from https://www.nitrc.org/projects/dtmframework/. This deformable template model (DTM) framework uses a progressive shape deformation technique based on a Laplacian surface representation of multi-level neighborhood triangular mesh vertices with flexible weight displacements. To build the DTM for right and left hippocampi we first co-register the hippocampal binary masks from all participants and generate the right and left hippocampal atlases by averaging the registered masks. Then, we apply marching cubes, and smooth and resample the triangular meshes of these atlases. The DTM is, then, non-rigidly deformed in a large-to-small scale to fit the individual shape of each hippocampus. This deformation process is iterative. At early iterations large deformations occur by propagating a external force that guides each vertex of the general model to the closest boundary across the surface. Throughout the iteration process, internal and external forces reach a balance while diminishing the rigidity at the neighborhood level so that the model deforms at smaller regions reproducing the local shape details. When this point is reached, we apply a rotation and scale-invariant transformation that limits the vertex transformations only to rotation, isotropic scale and translation to regularize the individual vertex transformations to those of the neighboring vertices using them as reference, avoiding that irregularities in the binary masks affect the fitting process.

### 2.3. Hippocampal shape analysis using an AI-based deformable mesh optimization framework for individual shape modeling

Our AI-based shape modeling method constructs the individual hippocampal shapes in a mesh representation by deforming the template mesh with a deep neural network. After the shape reconstruction, to derive the local deformity of individual hippocampal shape from the average template mesh for cross-sectional analysis, we apply the same overall procedure as (Hernández et al., 2017), which we use as baseline research.

The newly proposed shape modeling method consists hree stages as illustrated in Figure 1. In the first stage, prepare the target shape in a point cloud representation. As described in subsubsection 2.1.1, the binary masks of the hippocampi are generated either manually or semiautomatically, or using SynthSeg (Billot et al., 2023): a publicly available AI-based fully automated brain segmentation tool trained with synthetic data sampled from a generative model conditioned on the segmentations of the training set. Since manual masks have island artifacts along the boundary, we applied the same preprocessing procedure as the baseline method Hernández et al. (2017) to remove them. Holes and islands were eliminated through dilation and erosion using a 3 × 3 × 3 kernel, and the largest component was selected as the mask. From the boundary of the segmented binary masks, we extract the hippocampal point cloud. Subsequently, to avoid influencing clinical results by the individual brain size, the position of the point clouds is normalized using individuals’ intracranial volume (ICV). Specifically, the positions are scaled by dividing them by the cube root of the ICV. Finally, we derive the target shape, denoted as *Ptarget*, after registering onto the template mesh using the Iterative Closest Point (ICP) algorithm (Rusinkiewicz and Levoy, 2001).

**Figure 1:**
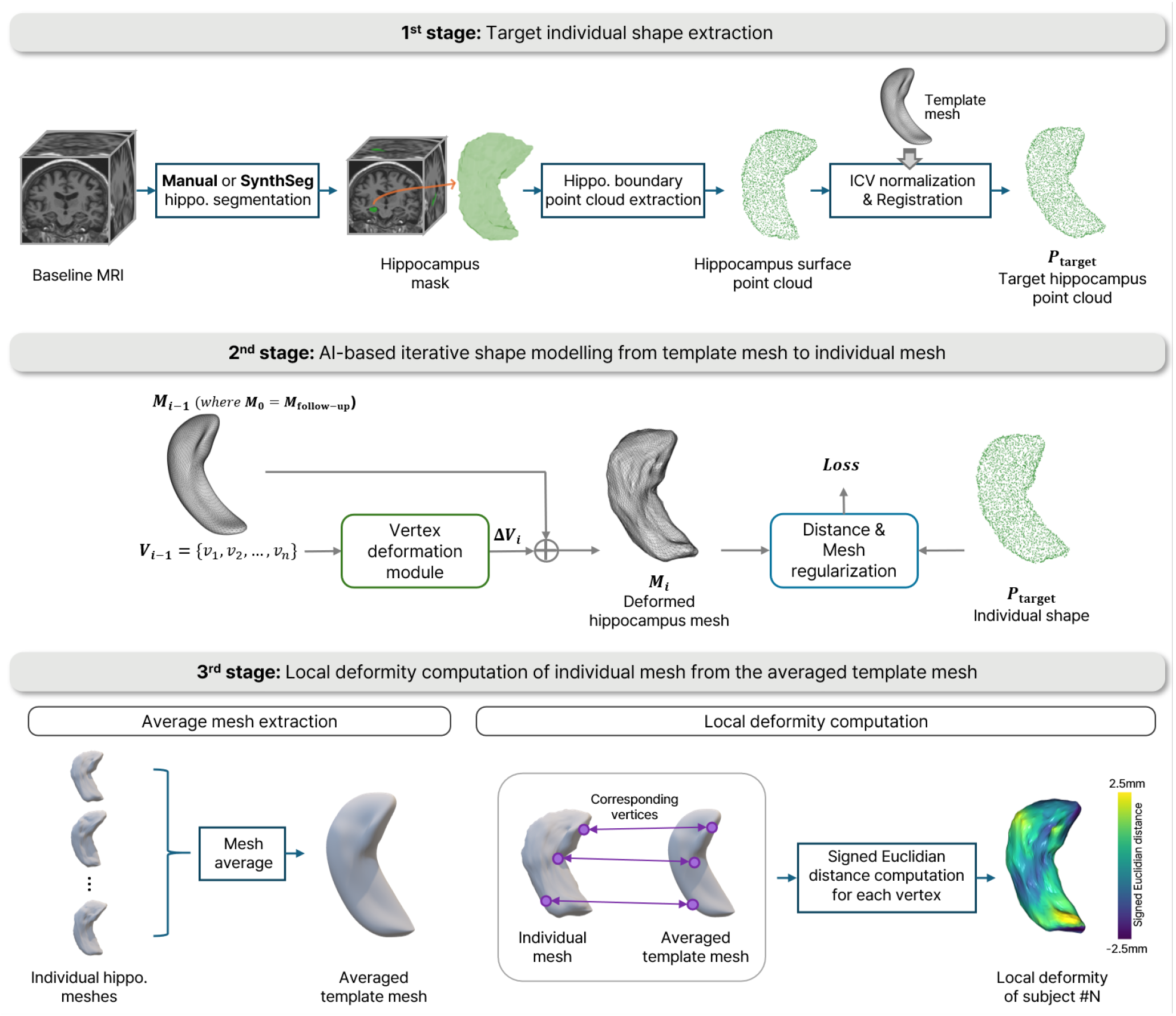
Overall pipelines to reconstruct the hippocampal shape from the template mesh and compute local deformity.

In the second stage, using the MLP-based PointNet architecture(Qi et al., 2017), the displacement of vertices is iteratively predicted and accumulated from the template mesh to align with the target point cloud. The iterative optimization scheme is based on Point2Mesh (Hanocka et al., 2020), which deforms the mesh while minimizing the loss function.

The objective function to reconstruct individual hippocampal meshes is the sum of two loss functions: a distance loss *Ldist* between the deformed mesh and the target point cloud, and a regularization loss *Lreg* for the deformed mesh.

The distance loss is:

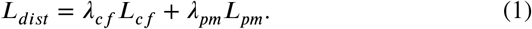

*Lcf* is the Chamfer distance between the selected points on the deformed mesh *Mi* and the point cloud *Pbaseline. Lpm* is a bidirectional distance between *Pbaseline* and the faces of *Mi*. The regularization loss is:

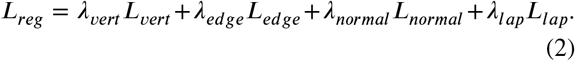

*Lvert* is the root mean square distance of the vertices moved, which induces the smallest movements as the deformable mesh approaches the target shape. *Ledge* is the variance of the edge lengths to prevent a skewed mesh. Normal consistency *Lnormal*, and Laplacian smoothness *Llap* are used to acquire a smooth deformed mesh.

We use the AdamW optimizer (Loshchilov and Hutter, 2017) for optimization with a learning rate of 5 × 10_^*−*4^_. The optimization is conducted over 5000 iterations with the learning rate halved every 1000 iterations. The template mesh consists of 4002 vertices, and the target point cloud contains 3000 points. For the Chamfer loss calculation, 3000 points are randomly selected from the deformed mesh. The hyperparameters {*λcf, λpm, λvert, λedge, λnormal, λlap*} are set to {0.5, 3, 1, 1500, 1, 5}.

In the third stage, we measure the local deformity of individual hippocampus from the average hippocampal template mesh. The average template mesh can be derived by averaging vertices from all optimized individual meshes. The average template mesh **M**_*avg* = {**v**_*avg*0, **v**_*avg*1, …, **v**_*avgn−*1} can be computed by averaging each vertex as follows where i-th individual mesh is

**M**_*i* = {**v**_*i*0, **v**_*i*1, …, **v**_*in−*1}:

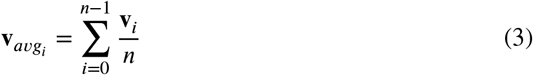

Then, we calculate the local deformity as the signed Euclidean distance between the vertices of the individual hippocampal meshes and the vertices of the average hippocampal mesh template. The signed Euclidean distance of each vertex of the i-th individual mesh is denoted as *LocalDef ormityi* = 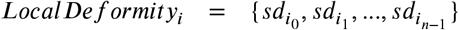 where each signed distance of the k-th vertex is compu_^*n*^_ted as follows:

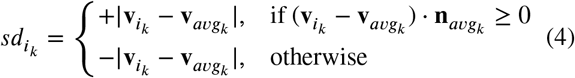

The 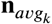 is the normal vector at the vertex 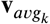. If the dot product o_^*k*^_f displacement of each vertex from the te_^*k*^_mplate vertex and the normal vector is larger than 0 which means that the displacement is above the tangent plane, the sign is positive, otherwise the sign is negative.

### 2.4. Experiments

We conducted eight experiments, listed in Table 1. The reference experiment listed as (0) reproduces the modeling work from the publication (Hernández et al., 2017), that uses the Laplacian-based deformation method as described in Section 2.2. It is applied to manually-edited hippocampal binary masks, and uses templates (i.e., one for right hippocampus and one for left hippocampus) obtained from 654 individuals with triangular meshes of 4002 vertices. Subsequent experiments are systematically designed to modify one of the following parameters: modeling method (experiments 1 onwards), segmentation method that generated the hippocampal binary masks (i.e., manual edit or SynthSeg described in subsubsection 2.1.1), number of individuals contributors to the average template (i.e., sample size of 654 or 601), vertex distribution across the shapes (i.e., uniform vs. non-uniform), and density of the template mesh reflected in the number of vertices (i.e., 4002 or 2001).

**Table 1.**
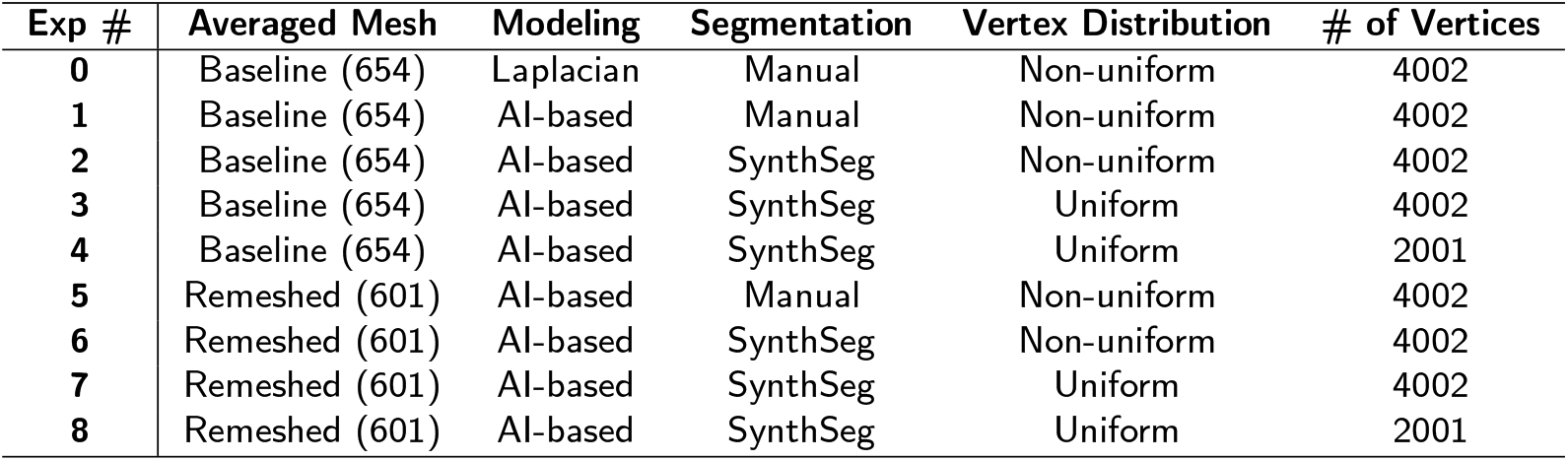
List of experiments exploring the relationship between hippocampal shape and cognition under varying conditions.

### 2.5. Statistical analyses

We explore how much cognitive functions in older age can be explained by local deformations, reproducing the statistical analyzes performed in (Hernández et al., 2017). For this, we use three general linear models with the deformation at each point of the hippocampal triangular meshes as the predictor (i.e., independent variable), each cognitive domain (i.e., *g-memory, g-speed* or *g*) as the response (i.e., dependent variable) for each model, and age, sex, and vascular risk score as covariates. The vascular risk score is an aggregate score of contemporaneous vascular risk derived from the presence/absence of old infarcts identified from the MRI scan, and the self-reported history of having had diabetes, hypertension, hypercholesterolemia, cardiovascular disease, and being currently, or just until recently, a smoker, which each participant provided during a medical interview. In our analyzes, we emphasize in the regions in which the associations between the hippocampal shape deformations with respect to the average template shape and the cognitive domains were statistically significant, with p-values less than 0.05. As shown in the overall results of experiments in Figure 2, the red color on the shapes indicates that the local position is significantly related to cognition.

**Figure 2:**
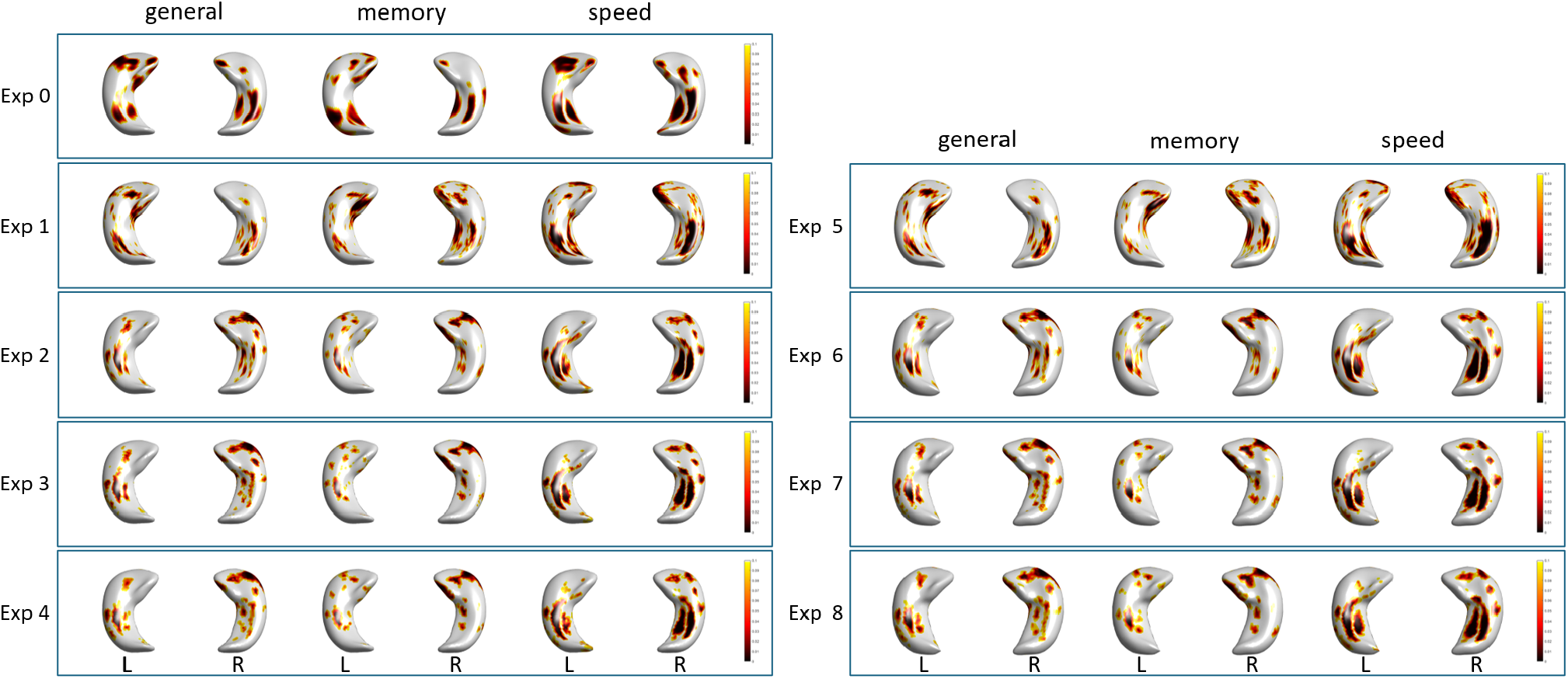
Overall results from each experiment. P-values from each statistical model. The local deformity on the regions with red color is significantly associated to the specific cognitive domain. Exp refers to the specific experiment, listed in Table 1.

### 2.6. Validation of the newly-proposed AI-based shape modeling method

We evaluated the performance of our AI-based shape modeling method with three spatial metrics: 1) the Dice Similarity Coefficient (DSC), 2) the average distance between the points of the individualized surface (i.e., mesh) models and the corresponding boundaries of the binary masks (in mm), and 3) the Hausdorff distance (in mm) also between the individual surfaces and the corresponding boundaries in binary masks.

## 3. Results

### 3.1. Laplacian-based shape modeling (Exp 0) vs. the AI-based shape modeling method proposed (Exp1)

We compared the clinically-relevant results from the Laplacian-based (Kim et al., 2019) (Exp 0) and our AI-based shape modeling methods (Exp 1) in the same 601 subjects, operating with hippocampal masks segmented manually/semi-automatically. The quality of surface reconstruction, the Laplacian-based method Hernández et al. (2017) reported an average Dice Similarity Coefficient (DSC) score of 0.96, with an interquartile range (IQR) of 0.027. Although the shape reconstruction in that study was manually adjusted to fit the target binary masks, our automated AI-based method obtained an improved mean DSC score of 0.97 with an IQR of 0.0131.

As shown in Figure 2, surface deformations in most ventral regions in the subiculum, pre-subiculum, CA2-3 and CA4 significantly associated with g and g-speed using the Laplacian-based model (Exp 0), were also associated using the AI-based modeling method (Exp 1). For g-memory, associations in these regions were replicated only for the right hippocampus. Exp 0 reported clusters of significant associations in the head and dorsal CA1 across the three domains, but these were very reduced in size or not visible in Exp 1. However, it must be noted that Hernández et al. (2017) (i.e., modeling reproduced in Exp 0) reported that after applying Type 1 error correction (false discovery rate) only associations with g-speed in the subiculum, presubiculum and ventral regions of body and tail and some reduced areas of CA1 remained significant. These regions were all statistically significantly associated with cognition using the AI-based modeling method (Exp 1).

Additionally, the AI-based shape modeling method (Exp 1) shows more sensitive patterns compared to the Laplacian-based method (Exp 0). This result is related to the algorithms of each method. The Laplacian-based method implies that certain vertices drags adjacent vertices together, making it prone to a blurring effect in clinically-relevant regions. On the other hand, the AI-based method deforms the vertices individually, allowing it to fit the target shape more accurately but revealing the clinically-relevant regions sensitively.

### 3.2. Manual (Exp1) vs. automatic (Exp2) segmentation masks

We explored how segmentation masks affect clinically-relevant outcomes in AI-based shape modeling through the analysis of results from Exp 1 and Exp 2. Manual segmentation is generally regarded as the gold standard. However, 3D manual segmentation presents significant challenges, as it is difficult to maintain consistent standards across images even for clinicians and well-trained image analysts and to achieve consistent masking throughout all three dimensions. With advances in deep learning, automatic segmentation tools have achieved near-perfect accuracy, leading to their growing adoption in medical image analysis. We use SynthSeg Billot et al. (2023), a deep learning-based brain segmentation tool, to generate hippocampal masks and compare them to manually created (or manually-corrected) masks in terms of clinically-relevant outcomes (i.e., associations between hippocampal shape and cognitive domains).

As seen in Exp 1 and Exp 2 of Figure 2, the regions significantly related to cognition vary depending on the segmentation masks used. Notably, the CA1 and part of the uncus in the head of the right hippocampus shows a strong relation with cognition (general, g-memory, and g-speed) when using SynthSeg masks (Exp 2) while the pattern is absent in the manual segmentations of Exp 0 and Exp 1.

We further investigate this discrepancy by comparing the manual and SynthSeg segmentations quantitatively and qualitatively. As shown in the plots in Figure 4, there is a considerable systematic difference in the number of hippocampal voxels masked by the two methods, with SynthSeg identifying around 1000 more voxels on average. Qualitatively, Figure 5(a) illustrates that SynthSeg covers a larger region. In addition, manual and manually rectified masks sometimes have island artifacts, since manual segmentation or rectification of automatically-generated masks is done on 2D slices (i.e., slice-by-slice), which leads to inconsistent boundaries when viewed in 3D. In contrast, SynthSeg produces clear and continuous boundaries without these artifacts. To investigate the larger voxel counts in SynthSeg, we examined the hippocampal mask boundaries. As can be seen from Figure 5(b), SynthSeg sometimes over-segments the voxels that appear to be cerebrospinal fluid (CSF) with near-black intensities in T1-weighted MRI as part of the hippocampus, and may include part of the amygdala.

**Figure 3:**
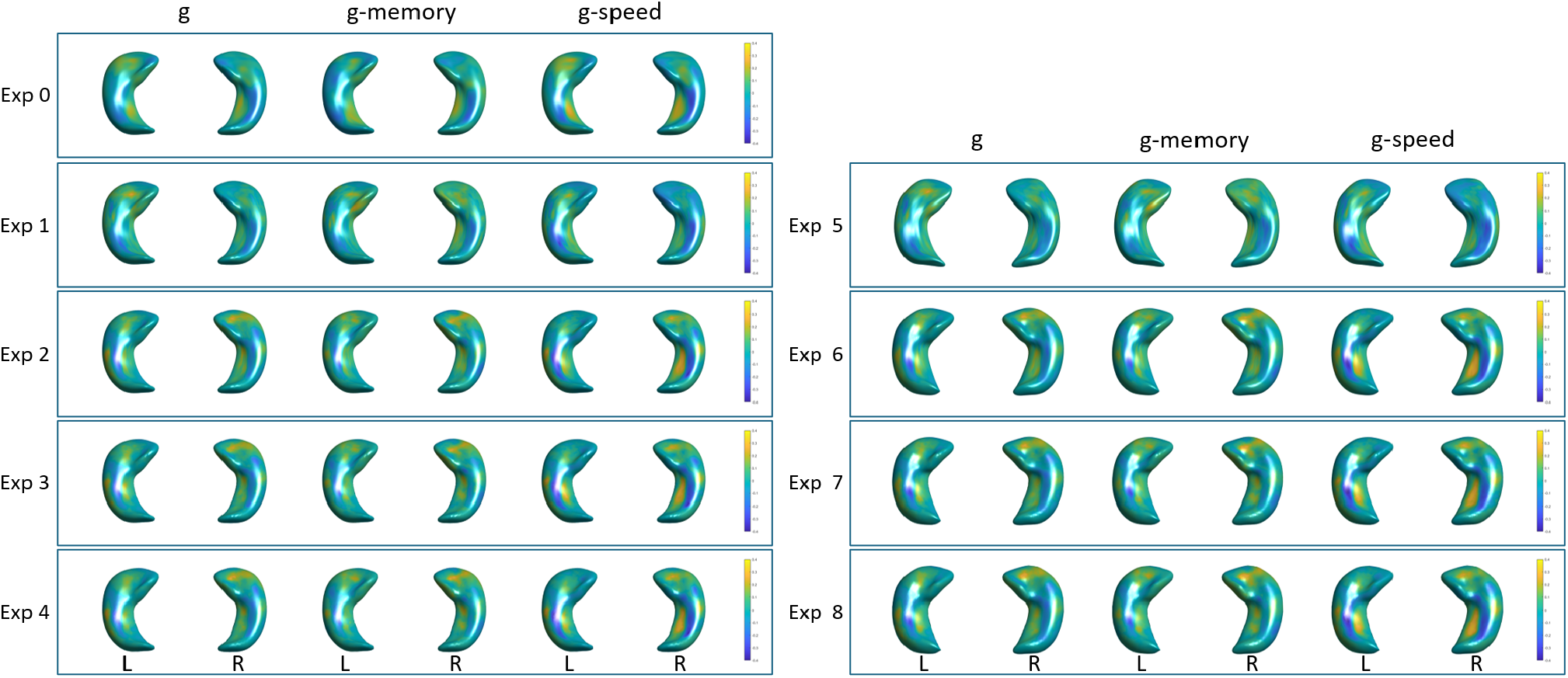
Overall results from each experiment. Beta coefficients from each model reflecting the magnitude of the association between the cognitive domain and the deformation in each point. Yellow areas represent positive associations (shape deformed outwards with respect to the average shape) while indigo areas represent negative associations (shape deformed inwards with respect to the average shape, i.e., atrophy)

**Figure 4:**
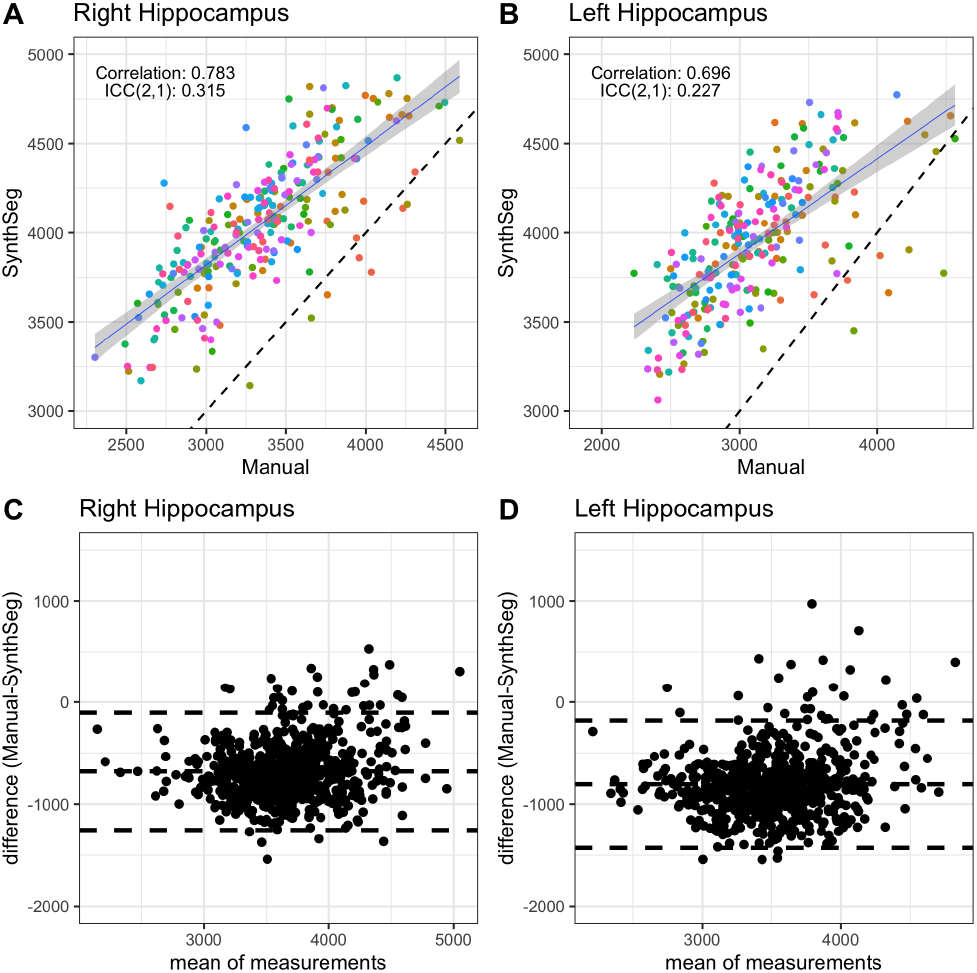
Quantitative comparison of manual and Synthseg segmentation based on the number of voxels that make up the volume of the region of interest.

**Figure 5:**
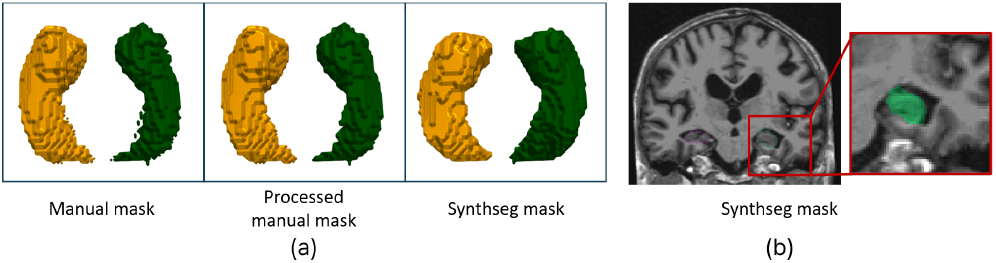
(a) Examples of segmentation masks on the same subject (b) Over-segmentation from Synthseg

We examined the impact of segmentation methods on the accuracy of shape modeling. Across all accuracy metrics, SynthSeg segmentation produced more accurate voxel-to-shape modeling results, as shown in Table 2. Additionally, SynthSeg-based shape modeling exhibited lower variance across all metrics, indicating greater consistency in the process. In contrast, the unclear boundaries of manual masks posed challenges, leading to less consistent shape modeling. In summary, segmentation methods can influence both clinical outcomes and the accuracy of the individual shape reconstruction. This highlights the importance of accurate segmentation for reliable clinical results.

**Table 2.** AI-based shape modeling accuracy under varied segmentation methods.

| Segmentation | Manual | SynthSeg |
| --- | --- | --- |
| DSC | 0.970 ± 0.026 | **0.978 ± 0.014** |
| Average dist. (mm) | 0.116 ± 0.098 | **0.090 ± 0.054** |
| Hausdorff dist. (mm) | 1.051 ± 0.287 | **1.024 ± 0.105** |

### 3.3. Use of different initial template meshes

The initial mesh is deformed to reconstruct individual hippocampal meshes as described in the second stage of Figure 1. In this section, we analyzed how the properties of the initial template mesh affect clinical results in AI-based shape modeling. Previous research (Hernández et al., 2017) utilized the initial template mesh shown in Figure 6(a). As shown in the figure, the vertex distribution in this mesh is non-uniform, leading to varying edge lengths across different regions of the hippocampus. Due to this non-uniformity, vertices in sparsely distributed areas have higher variance in deformation to capture the shapes of individual subjects. In contrast, vertices in densely distributed areas can cover individual meshes with less variance.

**Figure 6:**
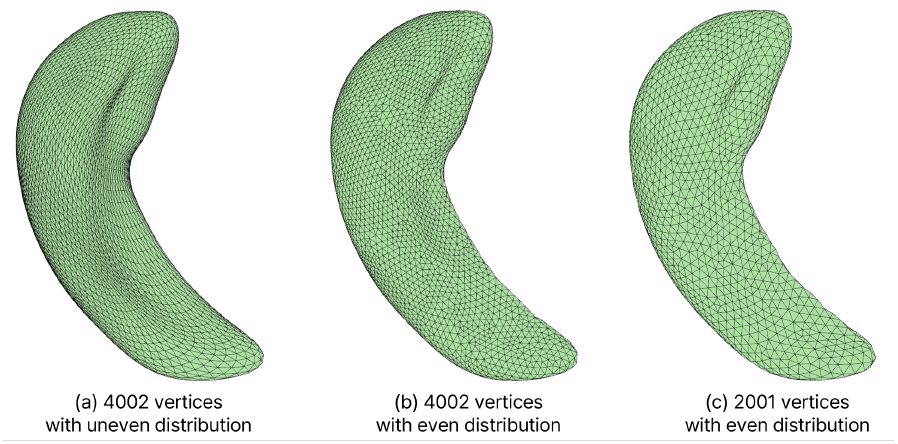
Initial template meshes for (a) Exp 0, 1, 2, 5, and 6 (b) Exp 3 and 7 (c) Exp 4 and 8.

Since the properties of the template mesh influence the construction of individual meshes, it is crucial to understand how the characteristics of the initial template mesh affect clinical outcomes. Therefore, in addition to the previously used template mesh (a) with non-uniformly distributed 4002 vertices, we created a new initial template mesh shown in (b). This new template mesh also has the same shape as the (a) and 4002 vertices but features a uniform vertex distribution. Furthermore, to evaluate the impact of vertex number, we analyze clinically-relevant results using another initial template mesh shown in (c), which has the same shape as (b) but 2001 uniformly distributed vertices (i.e., half of the number of vertices).

#### 3.3.1. Even vertices arrangement vs. uneven vertices arrangement

As shown in Figure 7, the variance in the local deformity at each vertex varies depending on the vertex distribution. In particular, the tail of the hippocampi (inferior point in the figure) with sparse vertices in the non-uniform template shows large variance. Despite these differences in variance of each vertex, the regions significantly associated with cognition, as seen in Exp 2 and Exp 3 of Figure 2 and Figure 3, remain similar. This suggests that the vertex distribution of the template mesh has minimal impact on these associations. Furthermore, it suggests that the AI-based shape modeling is robust to variations in vertex distribution..

**Figure 7:**
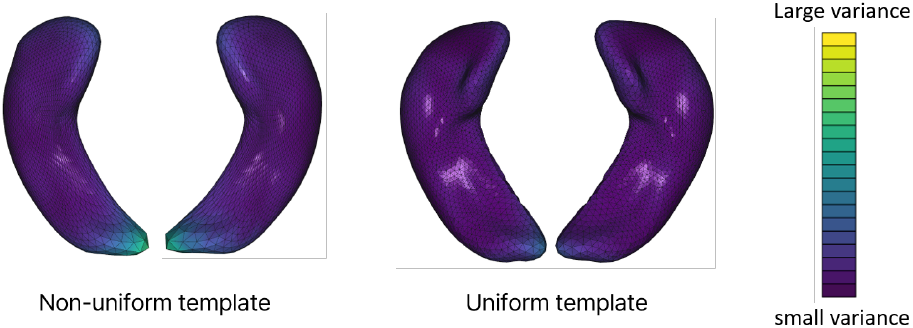
Variance in local deformity across subjects on templates with different vertex distributions.

#### 3.3.2. Different vertices number of template mesh

As shown in Exp 3 and Exp 4 of Figure 2 and Figure 3, the significantly associated regions and the magnitude of the associations remained similar regardless of vertex density. Although Exp 3 used 4002 vertices to reconstruct individual shapes, all regions displaying significant associations between cognitive domains and hippocampal shape local regions were reproduced in Exp 4, which used only 2001 vertices. This indicates that our AI-based shape modeling method consistently reconstructs shapes irrespective of the vertex density of the template meshes.

### 3.4. Results from using different averaged mesh

To compare the clinical results from our AI-based method with those from the baseline experiment (Exp 0) Hernández et al. (2017), we used the same average shape from Exp 0 for experiments Exp 1 through Exp 4. For experiments Exp 5 through Exp 8, we generated the average mesh by averaging the vertices of all individual meshes optimized in each experiment, as described in Equation 3. As shown in Figure 2, paired experiments using different average meshes (i.e. 1 & 5, 2 & 6, 3 & 7, 4 & 8) highlight similar significant regions on the hippocampi, suggesting that the choice of average mesh does not affect the relation between shape and cognition.

Meanwhile, the average mesh in Exp 5 shows a large discrepancy compared to Exp 6 while Exp 6, 7 and 8 have similar shapes. This indicates that the average shapes derived from manual vs SynthSeg segmentations differ significantly although they both use the same group of 601 subjects.

## 4. Discussion

We presented an AI-based hippocampal deformation model that optimizes the fitting of the vertices of the template mesh to a point cloud constructed from hippocampal binary masks using a deep neural network architecture. The newly presented AI-based method has higher sensitivity to small deformations (reflected in small average mesh-to-mask differences) and accuracy (reflected in the agreement between mesh and mask surfaces given by the DSC) than the traditional method with which it was compared. This high sensitivity and accuracy yields less statistically significant points that are associated with clinically relevant parameters as the samples involved in the analyzes are more heterogeneous. We compared the performance of this method with a state-of-the-art non-AI method in terms of its applicability to a clinically-relevant task: investigate the associations between hippocampal shape and cognitive domains.

Overall, the central area of the hippocampal body regions showing statistically significant associations between cognitive domains and hippocampal shape deformations with respect to a mean shape for cognitively normal older individuals at 72.6 years old using a Laplacian-based template deformation modeling principle and manually-edited binary masks, remain associated when an AI-based method using fully (i.e., also AI-based) automatically-generated binary masks are used, and the directions and magnitude of the associations are consistent throughout. However, associations of deformations in the head and extreme of the tail of the hippocampi with cognition differed between both methods in general. Hippocampal head and tail are always most affected by differences between segmentation methods, image quality, and point distribution in meshes. Thus, consequently, the shape deformation methods that are based on these segmented masks would yield contrasting results. This is not surprising, given that the different conventional methods that have previously analyzed the hippocampal shape deformations in relation to cognition, also yield inconsistent results precisely in these areas (Colom et al., 2013; Lim et al., 2012).

But importantly, this AI-based method consistently (i.e., across all experiments) revealed inward deformation in large areas of the hippocampal head -mainly for the right hippocampus- being associated with general memory. This is in agreement with findings from the large body of literature on hippocampal function that point at memory changes being associated with hippocampal shrinkage (S et al., 2020; Lim et al., 2012). Specifically in the CA1 head of the right hippocampus, the association was found in larger areas when using SynthSeg to generate the hippocampal masks, and the extent of these areas was not perceived to change with differences in the distribution (i.e., uniform vs. not uniform), numbers of mesh vertices and the template mesh used.

Despite the newly presented AI-based method being compared only with one of the traditional methods previously used by the community, the performance of this traditional method was compared already with two of the other most widely used non-AI hippocampal modeling methods: SPHARM-PDM (Styner et al. (2006) and LDDMM Raamana et al. (2014) yielding similar results Kim et al. (2014) and has been the base for recent developments Diers et al. (2023). AI-based modeling methods offer, in general, increased sensitivity compared with the traditional methods, where a deformation in a mesh vertex will affect those in the neighboring vertices prompting larger areas to be, consequently, affected, and offering smoother deformation patterns. But increased sensitivity does not come without a caveat: if binary masks are not carefully and accurately derived, or if the base MRI is noisy with artefacts causing imprecisions in the mask segmentation, the analysis using the AI-based modeling method can be misleading. Therefore, care must be taken in applying and interpreting the results from each method. As in other fields and applications, AI offers advantages in terms of precision and sensitivity for modeling hippocampal shape, but to capitalize in them, the whole pipeline, from image acquisition to statistical analyzes of the results, require careful design.

## Data Availability

The data used in and generated by this manuscript, and the LBC1936 study data are available by request from https://lothian-birth-cohorts.ed.ac.uk/data-access-collaboration

## Acknowledgments

The authors are grateful to LBC1936 participants for their dedication to the study. We are also grateful to the LBC research team who collected and collated the data used in this study, particularly Dr. Natalie A. Royle for visually inspecting and manually editing all hippocampal binary masks used in experiments 0, 1, and 5, the Edinburgh Clinical Research Facility nursing staff, and the radiographers of the Brain Research Imaging Centre.

This work was supported by Institute for Information & communications Technology Promotion(IITP) grant funded by the Korea government(MSIT) (No.00223446, Development of object-oriented synthetic data generation and evaluation methods). The LBC1936 study was supported by Age UK (the Disconnected Mind project), the UK Medical Research Council (MRC; G0701120, G1001245, MR/M013111/1, MR/R024065/1), joint funding from the Medical Research Council and the Biotechnology and Biological Sciences Research Council (MR/K026992/1 for the Centre for Cognitive Ageing and Cognitive Epidemiology), joint funding from the Biotechnology and Biological Sciences Research Council and the Economic and Social Research Council (BB/W008793/1), and the University of Edinburgh. MR Imaging and the statistical analyses presented here were further supported by the Row Fogo Charitable Trust (The Row Fogo Centre for Research into Ageing and the Brain; AD.ROW4.35. BRO-D.FID3668413), and UK Dementia Research Institute (Edin002, DRIEdi17/18, and MRC MC-PC-17113) which receives its funding from DRI Ltd, funded by the UK Medical Research Council. SRC was also supported by a Sir Henry Dale Fellowship jointly funded by the Wellcome Trust and the Royal Society (221890/Z/20/Z).

## References

Amunts, K., Kedo, O., Kindler, M., Pieperhoff, P., Mohlberg, H., Shah, N., Habel, U., Schneider, F., Zilles, K., 2005. Cytoarchitectonic mapping of the human amygdala, hippocampal region and entorhinal cortex: intersubject variability and probability maps. Anatomy and embryology 210, 343–352.

Anblagan, D., Valdés Hernández, M.C., Ritchie, S.J., Aribisala, B.S., Royle, N.A., Hamilton, I.F., Cox, S.R., Gow, A.J., Pattie, A., Corley, J., et al., 2018. Coupled changes in hippocampal structure and cognitive ability in later life. Brain and behavior 8, e00838.

Armitage, P., Chappell, F., MacLullich, A., Shenkin, S., Wardlaw, J.M., 2024. Normal reference T1-weighted MR images for the brain at ages 65-70 and 75-80 years, [dataset]. University of Edinburgh. Brain Research Imaging Centre. Centre for Clinical Brain Sciences.

Barnes, J., Foster, J., Boyes, R.G., Pepple, T., Moore, E., Schott, J.M., Frost, C., Scahill, R.I., Fox, N.C., 2008. A comparison of methods for the automated calculation of volumes and atrophy rates in the hippocampus. Neuroimage 40, 1655–1671.

Billot, B., Greve, D.N., Puonti, O., Thielscher, A., Van Leemput, K., Fischl, B., Dalca, A.V., Iglesias, J.E., et al., 2023. Synthseg: Segmentation of brain mri scans of any contrast and resolution without retraining. Medical image analysis 86, 102789.

Bird, C.M., Burgess, N., 2008. The hippocampus and memory: insights from spatial processing. Nature reviews neuroscience 9, 182–194.

Bongratz, F., Rickmann, A.M., Wachinger, C., 2024. Neural deformation fields for template-based reconstruction of cortical surfaces from mri. Medical Image Analysis 93, 103093.

Bouix, S., Pruessner, J.C., Collins, D.L., Siddiqi, K., 2005. Hippocampal shape analysis using medial surfaces. Neuroimage 25, 1077–1089.

Brechbühler, C., Gerig, G., Kübler, O., 1995. Parametrization of closed surfaces for 3-d shape description. Computer vision and image understanding 61, 154–170.

Cates, J., Fletcher, P.T., Styner, M., Shenton, M., Whitaker, R., 2007. Shape modeling and analysis with entropy-based particle systems, in: Information Processing in Medical Imaging: 20th International Conference, IPMI 2007, Kerkrade, The Netherlands, July 2-6, 2007. Proceedings 20, Springer. pp. 333–345.

Chen, X., Ravikumar, N., Xia, Y., Attar, R., Diaz-Pinto, A., Piechnik, S.K., Neubauer, S., Petersen, S.E., Frangi, A.F., 2021. Shape registration with learned deformations for 3d shape reconstruction from sparse and incomplete point clouds. Medical image analysis 74, 102228.

Colom, R., Stein, J.L., Rajagopalan, P., Martínez, K., Hermel, D., Wang, Y., Álvarez-Linera, J., Burgaleta, M., Quiroga, M.Á., Shih, P.C., et al., 2013. Hippocampal structure and human cognition: Key role of spatial processing and evidence supporting the efficiency hypothesis in females. Intelligence 41, 129–140.

Csernansky, J.G., Wang, L., Swank, J., Miller, J.P., Gado, M., Mckeel, D., Miller, M.I., Morris, J.C., 2005. Preclinical detection of alzheimer’s disease: hippocampal shape and volume predict dementia onset in the elderly. Neuroimage 25, 783–792.

Davies, R.H., Twining, C.J., Allen, P.D., Cootes, T.F., Taylor, C.J., 2003. Shape discrimination in the hippocampus using an mdl model, in: Information Processing in Medical Imaging: 18th International Conference, IPMI 2003, Ambleside, UK, July 20-25, 2003. Proceedings 18, Springer. pp. 38–50.

Deary, I.J., Gow, A.J., Taylor, M.D., Corley, J., Brett, C., Wilson, V., Campbell, H., Whalley, L.J., Visscher, P.M., Porteous, D.J., et al., 2007. The lothian birth cohort 1936: a study to examine influences on cognitive ageing from age 11 to age 70 and beyond. BMC geriatrics 7, 1–12.

Diers, K., Baumeister, H., Jessen, F., Düzel, E., Berron, D., Reuter, M., 2023. An automated, geometry-based method for hippocampal shape and thickness analysis. NeuroImage 276, 120182. URL: https://www.sciencedirect.com/science/article/pii/S1053811923003336, z10.1016/j.neuroimage.2023.120182.

García-Portugués, E., Meilán-Vila, A., 2023. Hippocampus shape analysis via skeletal models and kernel smoothing, in: Statistical Methods at the Forefront of Biomedical Advances. Springer, pp. 63–82.

Gerig, G., Muller, K.E., Kistner, E.O., Chi, Y.Y., Chakos, M., Styner, M., Lieberman, J.A., 2003. Age and treatment related local hippocampal changes in schizophrenia explained by a novel shape analysis method, in: Medical Image Computing and Computer-Assisted Intervention-MICCAI 2003: 6th International Conference, Montréal, Canada, November 15-18, 2003. Proceedings 6, Springer. pp. 653–660.

Hanocka, R., Metzer, G., Giryes, R., Cohen-Or, D., 2020. Point2mesh: a self-prior for deformable meshes. ACM Trans. Graph. 39.

Hasboun, D., Chantôme, M., Zouaoui, A., Sahel, M., Deladoeuille, M., Sourour, N., Duyme, M., Baulac, M., Marsault, C., Dormont, D., 1996. Mr determination of hippocampal volume: comparison of three methods. American journal of neuroradiology 17, 1091–1098.

Hernández, M.d.C.V., Cox, S.R., Kim, J., Royle, N.A., Maniega, S.M., Gow, A.J., Anblagan, D., Bastin, M.E., Park, J., Starr, J.M., et al., 2017. Hippocampal morphology and cognitive functions in community-dwelling older people: the lothian birth cohort 1936. Neurobiology of Aging 52, 1–11.

Kim, H., Park, J., 2024. Vertebral segmentation without training using differentiable appearance modeling of a deformable spine template, in: Medical Imaging 2024: Image Processing, SPIE. pp. 651–657.

Kim, J., Hernández, M.d.C.V., Park, J., 2019. Three-dimensional shape modeling and analysis of brain structures. JoVE (Journal of Visualized Experiments), e59172.

Kim, J., Valdes-Hernandez, M.d.C., Royle, N.A., Park, J., 2014. Hippocampal shape modeling based on a progressive template surface deformation and its verification. IEEE transactions on medical imaging 34, 1242–1261.

Kong, F., Wilson, N., Shadden, S., 2021. A deep-learning approach for direct whole-heart mesh reconstruction. Medical image analysis 74, 102222.

Lim, H.K., et al., 2012. Relationships between hippocampal shape and cognitive performances in drug-naïve patients with alzheimer’s disease. Neuroscience Letters 516, 124–129. URL: https://www.sciencedirect.com/science/article/pii/S0304394012004703, 10.1016/j.neulet.2012.03.072.

Loshchilov, I., Hutter, F., 2017. Decoupled weight decay regularization. arXiv preprint 1711.05101.

MacLullich, A., Ferguson, K., Deary, I., Seckl, J., Starr, J., Wardlaw, J., 2002. Intracranial capacity and brain volumes are associated with cognition in healthy elderly men. Neurology 59, 169–174.

Maruszak, A., Thuret, S., 2014. Why looking at the whole hippocampus is not enough—a critical role for anteroposterior axis, subfield and activation analyses to enhance predictive value of hippocampal changes for alzheimer’s disease diagnosis. Frontiers in cellular neuroscience 8, 95.

Mueller, S.G., Schuff, N., Yaffe, K., Madison, C., Miller, B., Weiner, M.W., 2010. Hippocampal atrophy patterns in mild cognitive impairment and alzheimer’s disease. Human brain mapping 31, 1339–1347.

Mulder, E.R., de Jong, R.A., Knol, D.L., van Schijndel, R.A., Cover, K.S., Visser, P.J., Barkhof, F., Vrenken, H., Initiative, A.D.N., et al., 2014. Hippocampal volume change measurement: quantitative assessment of the reproducibility of expert manual outlining and the automated methods freesurfer and first. Neuroimage 92, 169–181.

Papp, K.V., Kaplan, R.F., Springate, B., Moscufo, N., Wakefield, D.B., Guttmann, C.R., Wolfson, L., 2014. Processing speed in normal aging: effects of white matter hyperintensities and hippocampal volume loss. Aging, Neuropsychology, and Cognition 21, 197–213.

Patenaude, B., Smith, S.M., Kennedy, D.N., Jenkinson, M., 2011. A bayesian model of shape and appearance for subcortical brain segmentation. Neuroimage 56, 907–922.

Qi, C.R., Su, H., Mo, K., Guibas, L.J., 2017. Pointnet: Deep learning on point sets for 3d classification and segmentation, in: Proceedings of the IEEE conference on computer vision and pattern recognition, pp. 652–660.

Raamana, P.R., Rosen, H., Miller, B., Weiner, M.W., Wang, L., Beg, M.F., 2014. Three-class differential diagnosis among alzheimer disease, frontotemporal dementia, and controls. Frontiers in neurology 5, 71.

Reuben, A., Brickman, A.M., Muraskin, J., Steffener, J., Stern, Y., 2011. Hippocampal atrophy relates to fluid intelligence decline in the elderly. Journal of the International Neuropsychological Society 17, 56–61.

Rusinkiewicz, S., Levoy, M., 2001. Efficient variants of the icp algorithm, in: Proceedings third international conference on 3-D digital imaging and modeling, IEEE. pp. 145–152.

s, P.T., et al., 2020. Hippocampal shape is associated with memory deficits in temporal lobe epilepsy. Annals of neurology 88, 170–182.

Scheltens, P., Leys, D., Barkhof, F., Huglo, D., Weinstein, H., Vermersch, P., Kuiper, M., Steinling, M., Wolters, E.C., Valk, J., 1992. Atrophy of medial temporal lobes on mri in” probable” alzheimer’s disease and normal ageing: diagnostic value and neuropsychological correlates. Journal of Neurology, Neurosurgery & Psychiatry 55, 967–972.

Scher, A.I., Xu, Y., Korf, E., White, L.R., Scheltens, P., Toga, A.W., Thompson, P.M., Hartley, S., Witter, M., Valentino, D.J., et al., 2007. Hippocampal shape analysis in alzheimer’s disease: a population-based study. Neuroimage 36, 8–18.

Shen, D., Moffat, S., Resnick, S.M., Davatzikos, C., 2002. Measuring size and shape of the hippocampus in mr images using a deformable shape model. Neuroimage 15, 422–434.

Shen, L., Ford, J., Makedon, F., Saykin, A., 2003. Hippocampal shape analysis: surface-based representation and classification, in: Medical Imaging 2003: Image Processing, SPIE. pp. 253–264.

Styner, M., Gerig, G., Lieberman, J., Jones, D., Weinberger, D., 2003. Statistical shape analysis of neuroanatomical structures based on medial models. Medical image analysis 7, 207–220.

Styner, M., Oguz, I., Xu, S., Brechbühler, C., Pantazis, D., Levitt, J.J., Shenton, M.E., Gerig, G., 2006. Framework for the statistical shape analysis of brain structures using spharm-pdm. The insight journal, 242.

Van Strien, N.M., Widerøe, M., Van De Berg, W.D., Uylings, H.B., 2012. Imaging hippocampal subregions with in vivo mri: advances and limitations. Nature Reviews Neuroscience 13, 70–70.

Voineskos, A.N., Winterburn, J.L., Felsky, D., Pipitone, J., Rajji, T.K., Mulsant, B.H., Chakravarty, M.M., 2015. Hippocampal (subfield) volume and shape in relation to cognitive performance across the adult lifespan. Human brain mapping 36, 3020–3037.

Wang, N., Zhang, Y., Li, Z., Fu, Y., Liu, W., Jiang, Y.G., 2018. Pixel2mesh: Generating 3d mesh models from single rgb images, in: Proceedings of the European conference on computer vision (ECCV), pp. 52–67.

Wardlaw, J.M., Bastin, M.E., Valdés Hernández, M.C., Maniega, S.M., Royle, N.A., Morris, Z., Clayden, J.D., Sandeman, E.M., Eadie, E., Murray, C., et al., 2011. Brain aging, cognition in youth and old age and vascular disease in the lothian birth cohort 1936: rationale, design and methodology of the imaging protocol. International Journal of Stroke 6, 547–559.

Wechsler, D., 1997. Wechsler adult intelligence scale-iii. Frontiers in Psychology.

Wechsler, D., 1998. WMS-III (UK) Administration and Scoring Manual. 3rd edn (The Psychological Corporation: London. UK.

Wickramasinghe, U., Remelli, E., Knott, G., Fua, P., 2020. Voxel2mesh: 3d mesh model generation from volumetric data, in: Medical Image Computing and Computer Assisted Intervention–MICCAI 2020: 23rd International Conference, Lima, Peru, October 4–8, 2020, Proceedings, Part IV 23, Springer. pp. 299–308.

Woolard, A.A., Heckers, S., 2012. Anatomical and functional correlates of human hippocampal volume asymmetry. Psychiatry Research: Neuroimaging 201, 48–53.

Yushkevich, P.A., Pluta, J.B., Wang, H., Xie, L., Ding, S.L., Gertje, E.C., Mancuso, L., Kliot, D., Das, S.R., Wolk, D.A., 2015. Automated volumetry and regional thickness analysis of hippocampal subfields and medial temporal cortical structures in mild cognitive impairment. Human brain mapping 36, 258–287.

